# Exploring the barriers and enablers of oral health care utilization and safe oral sex behaviour among Transgender women of Malaysia: a qualitative study

**DOI:** 10.1101/2024.04.10.24305651

**Authors:** Lahari A Telang, Abdul Rashid, Aoife G Cotter

## Abstract

**Background:** Transgender women in Malaysia are vulnerable and marginalized. They experience unique social and interpersonal challenges that contribute to relatively unmet health care needs thereby increasing their risk of acquiring HIV and sexually transmitted infections (STIs). With research pertaining to oral health of transgender women being sparse in literature, this study aimed to understand their experiences by exploring the barriers and enablers of oral health care utilization as well as safe sexual behaviour relating to oral transmission of STIs among Transgender women.

**Methods:** Semi structured in-depth interviews (n=20) with a group of urban dwelling transgender women (mean age= 39.8 years) in Northern Malaysia and Focus group discussion (n=7) with key informants was conducted to gain insights into the needs of the community. Participants were recruited through a snow-balling method of sampling with the help of transgender women community workers. The data obtained was coded and transcribed and subjected to thematic analysis to interpret and derive major themes and subthemes. Community advocates were involved in planning of the study.

**Results:** All of participants reported experiencing high levels of stigma and discrimination in daily life. Routine engagement in oral sex practices (100%, n=20) was reported with multiple partners (60%, n=12) with a perceived low level of risk of transmission of STIs through this practice (80%, n=16). Nighty percent (n=18) never used barriers or inconsistently used them during oral sexual practices. Additionally, low utilization of dental services was reported with 80% (n=16) not having visited a dentist in the past 12 months or seeking self-medication or unprofessional care. The themes that emerged from the qualitative analysis were key to understanding the experiences of the community.

**Conclusion:** The results identified gaps in awareness of oral transmission of STIs among the transgender women. The importance of social support in positively influencing health promotion as well as improving health care and dental care utilization was highlighted. The development of an educational intervention is proposed as an instrument to address these gaps and provide support.

## BACKGROUND

Transgender women are individuals who are assigned male sex at birth but identify as female.(1, 2) In Malaysia, transgender women face stigma and discrimination and are subject to criminal prosecution by both civil and religious laws of the country. The inability to express their identities freely leads to transgender women being marginalized by society and by choice. Such marginalization may push them to sex work as a means of earning a living.(3-5) Health and wellbeing takes a toll as the basic determinants of health such as social security, education, employment, family support and health care are undermined. (6, 7)

Malaysia is an upper-middle income country and is one of a few countries in Asia that has achieved Universal Healthcare Utilization (UHC).(8) This means access to health care services including sexual, mental and oral health is ensured to all citizens without exposing the user to financial hardship (9, 10). Although healthcare is accessible for all, it fails to address the multiple healthcare needs of transgender women. This could be because of the poor cultural acceptance of transgender persons by the community including health care providers and also by the transgender woman herself who feels she is stigmatized and refuses treatment for fear of facing discrimination. This creates barriers to rightful utilization of health care services.(6, 11)

The Ministry of Health (MoH) Malaysia recognises transgender women as one of the key population groups who are vulnerable, and have a high prevalence of HIV and STIs (National Strategic Plan for Ending AIDS NSPEA 2016-2030) (2, 12) The estimates of transgender women living with HIV was reported to be 10.9% in 2017 (13) and 12.4% in 2018 among transgender women sex workers, along with high rates of syphilis reported.(14) Even though HIV/AIDS knowledge was noted to be adequate among transgender women, it did not necessarily translate into safe sexual practices or better attitude towards HIV.(15) Condom usage was found to be inconsistent even when the knowledge was adequate, and the attitude of transgender women was not found to significantly predict HIV related high risk behaviour. (15) Significant gaps have been highlighted in empirical data required for the scale-up of HIV prevention programs such as treatment uptake, adherence and HIV continuum of care, especially for a developing country like Malaysia (16) (17, 18).(19) Globally, public health intervention strategies to create awareness and curb the spread of sexually transmitted infections such as community education through key stake holders, distribution of free test kits and door to door visits and newer technology based intervention have been recommended as promising methods of reaching the target population.(20, 21)

With HIV taking the centre stage, the emphasis on prevention is directed towards spread of infection through anal sex for both men who have sex with men and transgender women.(22, 23) Even when it is well recognised that transmission of STI causing pathogens, both viral and bacterial, occurs during oral sexual activities, less emphasis is placed on oral sex as a risky sexual behaviour (22-24). This may be due to the fact that STIs in the oral and oral pharyngeal region are mostly asymptomatic, with manifestations seen less often in the anterior part of oral cavity and causing only an occasional sore throat when harboured in the tonsil or pharynx. (23, 25). The health of the oral cavity can have a direct impact on the transmission of infections. Unhealthy periodontal conditions, open sores or ulcers on the oral mucosa and bleeding from the gums are known to increase the risk of transmission.(26) According to the World Health Organization (WHO), disproportionately higher burden of oral diseases effect the most vulnerable and disadvantaged populations of the world.(27) Transwomen as a community, may present with unique oral health care needs in relation to access and utilization of services. There is a general lack of evidence-based research in this regards; with health disparities and oral health care needs of this population being relatively unmet globally. (7) Professional education regarding oral health and oral health disparities of individuals identified as gender minorities is also found to be lacking globally.(9)(10)

It has been recommended that for any healthcare intervention strategies to be effective they must be culturally sensitive and tailor made to the target population (21, 28). However in the case of transgender women, studies have shown that they are often subsumed in interventions with other key populations such as men who have sex with men which may lead to more generalized results that are not exclusive to transgender women. (29, 30) Thus the current study was designed exclusively for transgender women of Malaysia with an aim to identify their oral health care needs, understand their experiences by exploring their behavioural characteristics that pose as barriers and enablers of oral health care utilization as well as safe sexual behaviour relating to oral transmission of STIs.

## METHODOLOGY

The study was undertaken as a preliminary part of a larger study looking into oral health and sexual health relating to oral STIs of transgender women in Malaysia. Qualitative analysis methodology was adopted to gain depth in understanding and uncover nuances and meaning to this exploratory research.

### Ethics

The study received ethics approval by the University College Dublin, UCD Human Research Ethics Committee – Sciences (HREC-LS) (LS-C-22-158-Telang-Cotter) and the Joint Penang Independent Ethics Committee (JPEC 22-0002). The participant information sheet and informed consent were made available in English as well as translated to local language (Bahasa Malaysia). Details of privacy and confidentiality of the participants were explained clearly. The participants were ensured that all communications shall remain professional at all times relating only to the research project.

The Health Belief Model (HBM)(31) and the Andersen’s Behaviour Model of health services use(32) were used as frameworks to devise the open ended questions guide for the semi-structured interviews (Table 1) and focus group discussion. Initial meetings between the principal author and few of the transgender women community advocates were done in informal settings to explore the general frame of mind of the community. The community advocates provided feedback regarding the study design and assisted with recruitment of participants. They not only supported the interpretation of the preliminary data but also the follow-up part of the study.

**Table 1:**
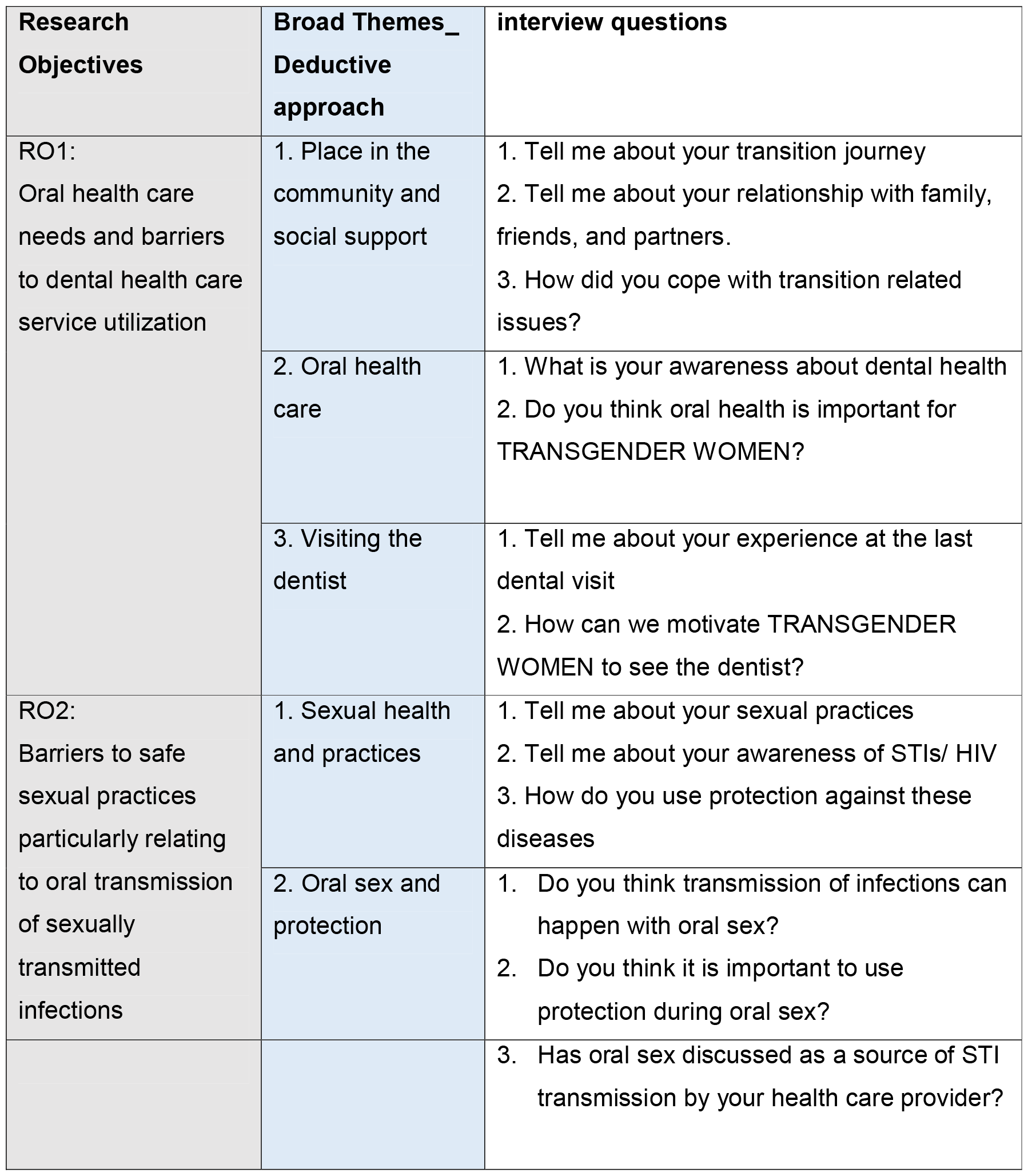
Guide to Semi-structured In-depth Interviews.

| **Research Objectives** | **Broad Themes_ Deductive approach** | **interview questions** |
| --- | --- | --- |
| RO1:<br>Oral health care needs and barriers to dental health care service utilization | 1. Place in the community and social support | 1. Tell me about your transition journey<br>2. Tell me about your relationship with family, friends, and partners.<br>3. How did you cope with transition related issues? |
|  | 2. Oral health care | 1. What is your awareness about dental health<br>2. Do you think oral health is important for TRANSGENDER WOMEN? |
|  | 3. Visiting the dentist | 1. Tell me about your experience at the last dental visit<br>2. How can we motivate TRANSGENDER WOMEN to see the dentist? |
| RO2:<br>Barriers to safe sexual practices particularly relating to oral transmission of sexually transmitted infections | 1. Sexual health and practices | 1. Tell me about your sexual practices<br>2. Tell me about your awareness of STIs/ HIV<br>3. How do you use protection against these diseases |
|  | 2. Oral sex and protection | 1. Do you think transmission of infections can happen with oral sex?<br>2. Do you think it is important to use protection during oral sex? |
|  |  | 3. Has oral sex discussed as a source of STI transmission by your health care provider? |

**Table 1:**
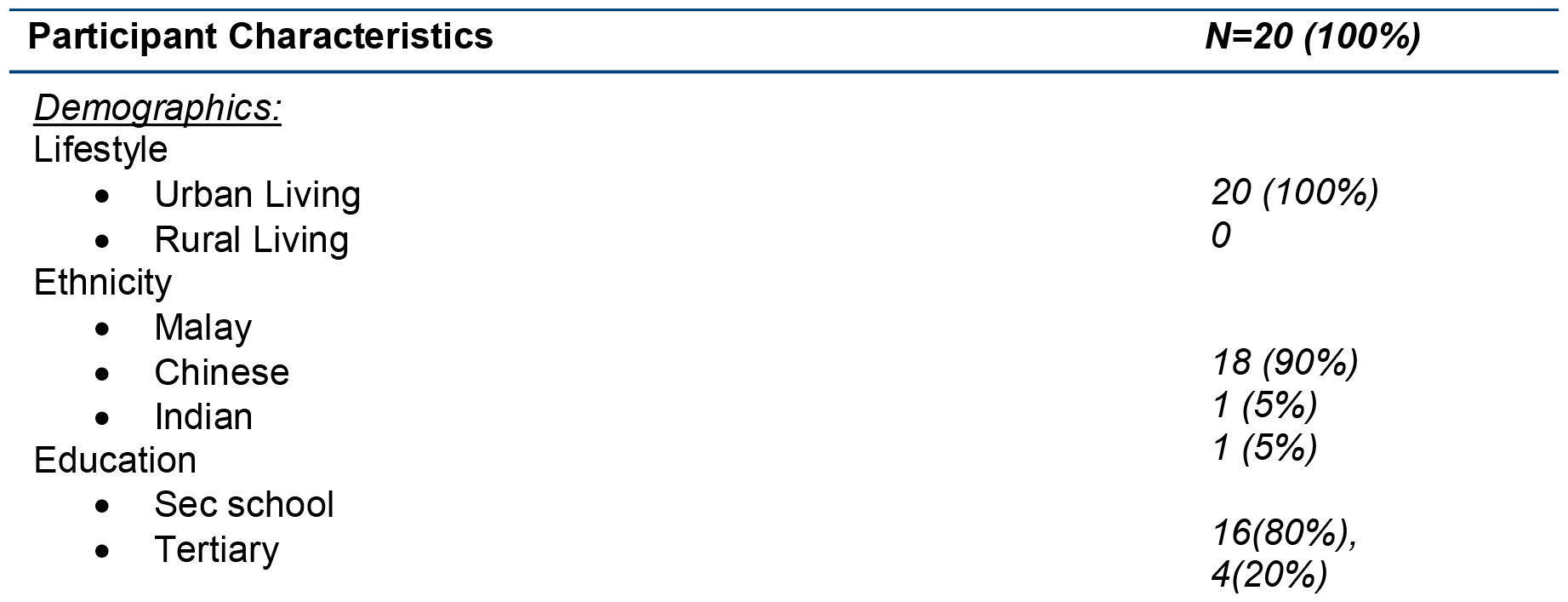
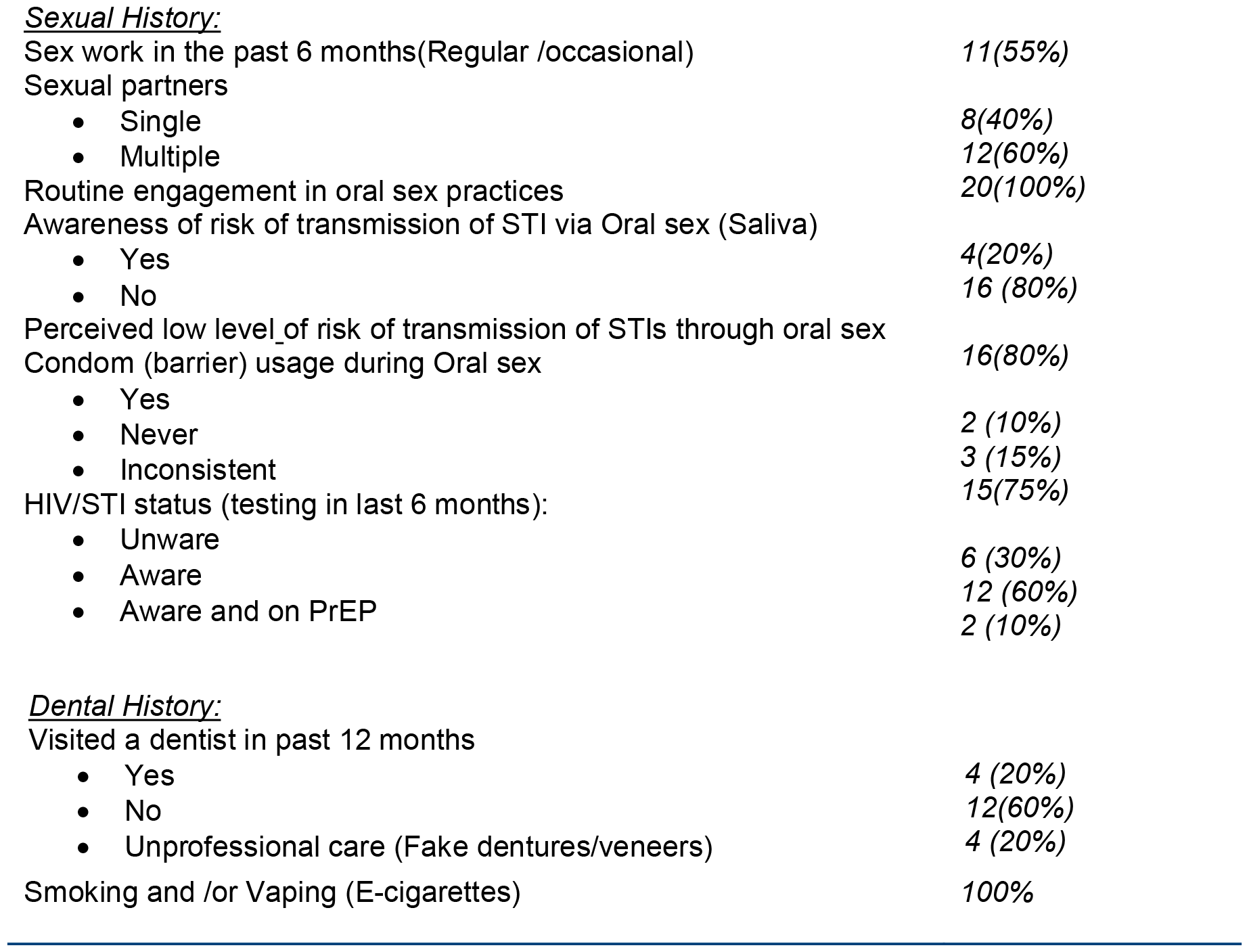
Participant demographics and history.

| **Participant Characteristics** | **N=20 (100%)** |
| --- | --- |
| <u>Demographics:</u> |  |
| Lifestyle |  |
| • Urban Living | 20 (100%) |
| • Rural Living | 0 |
| Ethnicity |  |
| • Malay | 18 (90%) |
| • Chinese | 1 (5%) |
| • Indian | 1 (5%) |
| Education |  |
| • Sec school | 16(80%), |
| • Tertiary | 4(20%) |
| Sexual History: |  |
| Sex work in the past 6 months(Regular /occasional) | 11(55%) |
| Sexual partners |  |
| • Single | 8(40%) |
| • Multiple | 12(60%) |
| Routine engagement in oral sex practices | 20(100%) |
| Awareness of risk of transmission of STI via Oral sex (Saliva) |  |
| • Yes | 4(20%) |
| • No | 16 (80%) |
| Perceived low level_of risk of transmission of STIs through oral sex | 16(80%) |
| Condom (barrier) usage during Oral sex |  |
| • Yes | 2 (10%) |
| • Never | 3 (15%) |
| • Inconsistent | 15(75%) |
| HIV/STI status (testing in last 6 months): |  |
| • Unware | 6 (30%) |
| • Aware | 12 (60%) |
| • Aware and on PrEP | 2 (10%) |
| Dental History: |  |
| Visited a dentist in past 12 months |  |
| • Yes | 4 (20%) |
| • No | 12(60%) |
| • Unprofessional care (Fake dentures/veneers) | 4 (20%) |
| Smoking and /or Vaping (E-cigarettes) | 100% |

### In-depth interviews

Initial participants were referred by the local transgender women community advocates. From there on the other participants were recruited through word of mouth using a snow-balling method of sampling with the help of the initial participants. Invitations for participation were sent out via personal text messages. Any individual identifying oneself as a transgender woman who were able to give informed consent, aged 18 years and above, with a minimum education level of primary school of grade 6 and currently residing in Malaysia were invited to participate in the study. Other inclusion criteria included participants who were currently sexually active, participants who may or may not be co-infected with HIV and/or other STIs, and current smart phone users.

Written informed consent was obtained and semi structured in-depth interviews were conducted (n=20). Face to face interviews were conducted for majority of the participants (n=16), with only two interviews done as group interviews with two participants together at each interview (n=4).This was done to accommodate the participants’ personal choices and preferences. All interviews were held at a neutral space based on the participant’s convenience and were audio recorded using an audio recorder. The interviews lasted from 30 minutes to 90 minutes duration. The interviews were conducted by the principal author and a trained research assistant who is a transgender woman with experience in transgender health outreach and advocacy. Both English and Bahasa Malaysia were used in the interviews and the help of the research assistant for the project was sought wherever deemed necessary to facilitate better understanding of variations in the local dialects and cultural nuances. The interview guide questions were flexible used to initiate the conversation and understand the participants views on each of the aspects listed in the guide (Table 1). Documentation of non-verbal cues during the interview was made using hand written notes and references were drawn after the interview by the main investigator. Saturation in data was determined as the point after which no new information was obtained and any further data collection through subsequent interviews would be redundant.(33) During the course of the study period participants identified with immediate health care needs or deemed to be at risk were intended to be referred to the nearest public health clinic or physician of participant’s choice and/or for psychological counselling. However, none of the participants in the current study required any of these services at the time of the interview. No participants withdrew after providing consent.

### Focus group discussion

The preliminary data from the interviews revealed key insights into the needs of the community and broad perspectives of the barriers and facilitators of oral and sexual health care. To a get a more detailed understanding of the situation, multiple stakeholder feedback was deemed essential. This was important to consolidate and validate the themes that were derived from the interviews. Hence a focus group discussion with a diverse group of participants was performed. The participants (n=7) were recruited through purposeful sampling. The group included members who would contribute significantly to the discussion and were directly or indirectly involved with transgender women health care in Malaysia. Two transgender women community advocates, an activist identifying as a transgender woman, an artist identifying as a transgender woman, a physician from the local STI “friendly clinic”, a psychological counsellor specializing in gender and sexual minorities’ matters and a general dentist who was working for a local non-governmental organization (NGO) for women’s welfare and vulnerable groups.

The discussion lasted for over two and half hours with each of the participants validating the concerns raised through the codes generated from the interviews. They also contributed information regarding the current position of transgender women in society, safe sexual practices and health care utilization. Through this discussion the suitability of an educational intervention to increase awareness was explored.

### Coding and data analysis

The interview data was then transcribed and coding was done using Nvivo 12 software. A multistep process of coding was done. The initial codes generated manually were compared with the ones generated using the software and a consensus was reached by the investigators. The most important ideas that emerged were coded and similar codes were clustered into minor themes. Thematic analysis was used to interpret and derive major themes.(34) Direct quotations from participants about their experiences, opinions, knowledge and feelings were also recorded as data. Methodological orientation of the analysis was guided by principles of Grounded Theory& Narrative inquiry.(35) A similar process of data coding and analysis was adopted for the focus group discussion data as well. Separate set of minor themes and major themes were derived from the data.

## RESULTS

The demographics and history of in depth interview participants are outlined in table 2. All participants of the in depth interviews identified as transgender women (n=20) with a median age of 39 years (27-58 years). All of them reported experiencing high levels of stigma and discrimination at some point in daily life. Majority had secondary school level of education (n=16(80%)), with a few of them receiving tertiary education (n=4 (20%)). While all of them were living in urban societies, 6(30%) of them engaged in sex work for a living. Only 2(10%) had ever used pre-exposure prophylaxis (PrEP). Routine engagement in oral sex practices *(20(100%))* was reported with multiple partners *(12(60%))* with a perceived low level of risk of transmission of STIs through this practice *(16(80%))*. Almost all (18(90%)) never used barriers or inconsistently used them during oral sexual practices. Additionally, low utilization of dental services was reported with the majority (16(80%)) not having visited a dentist in the past 12 months.

**Table 2:**
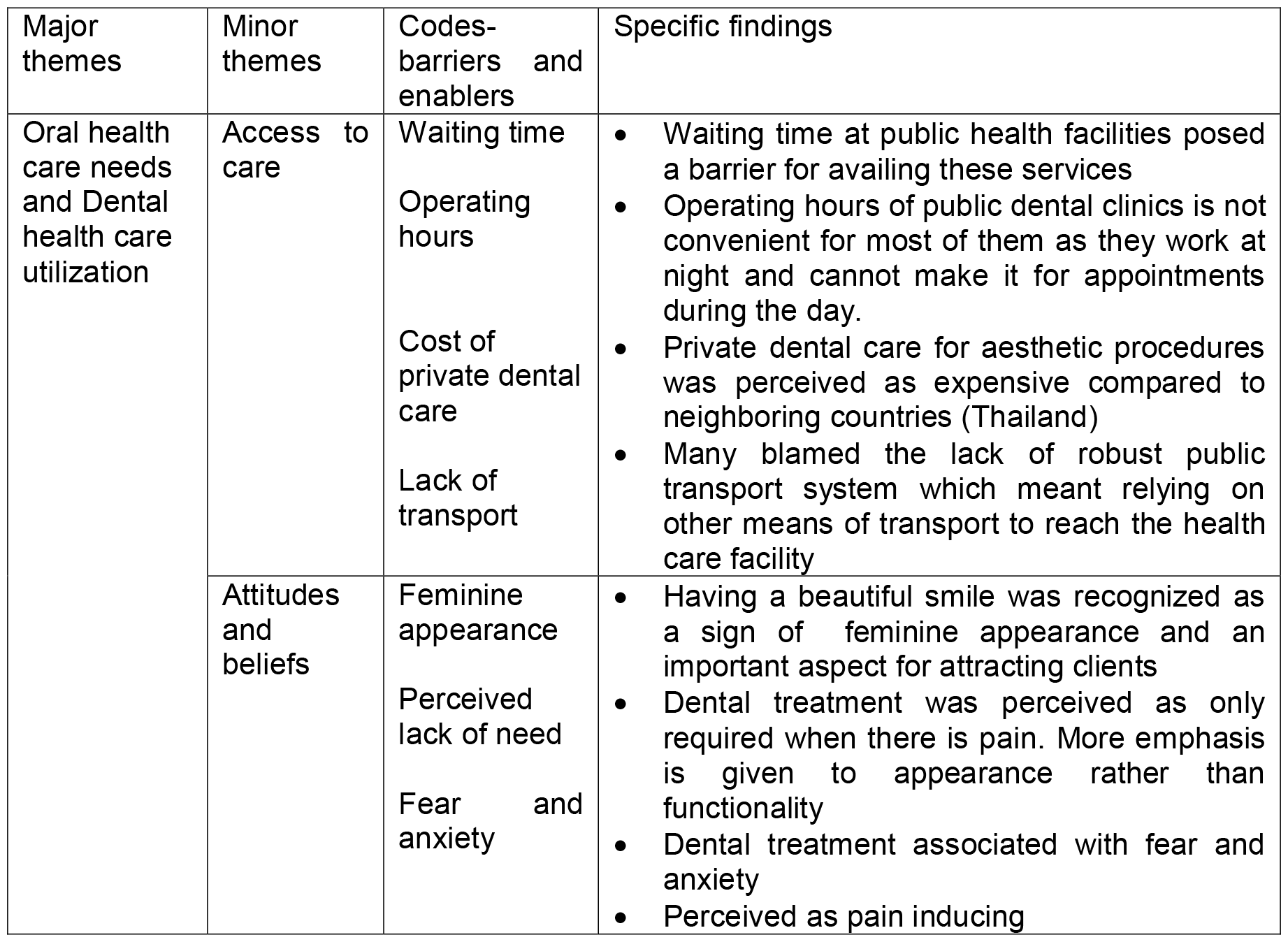
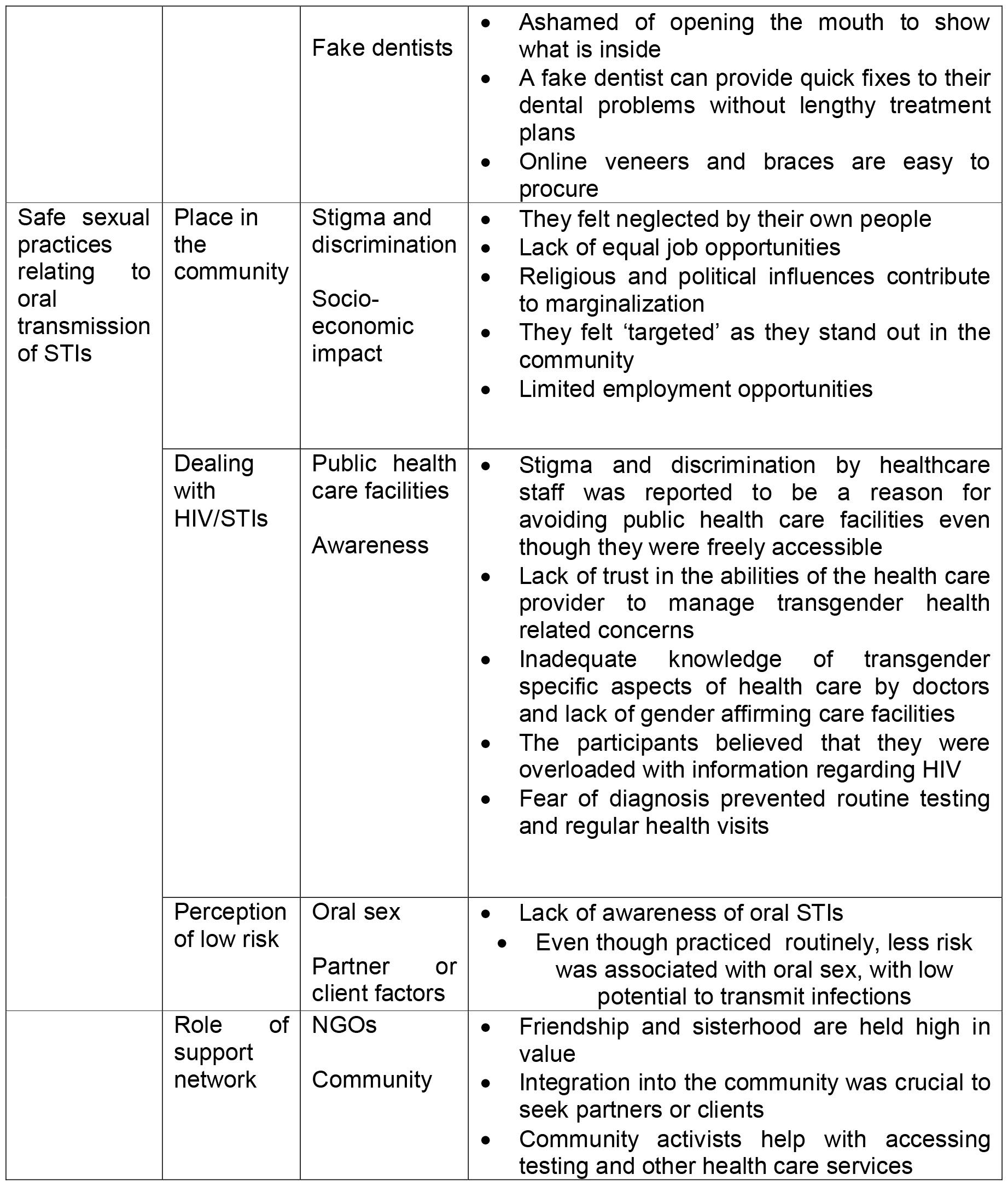
Summary of findings of thematic analysis:

| Major themes | Minor themes | Codes-barriers and enablers | Specific findings |
| --- | --- | --- | --- |
| Oral health care needs and Dental health care utilization | Access to care | Waiting time<br>Operating hours<br>Cost of private dental care<br>Lack of transport | Waiting time at public health facilities posed a barrier for availing these services<br>Operating hours of public dental clinics is not convenient for most of them as they work at night and cannot make it for appointments during the day.<br>Private dental care for aesthetic procedures was perceived as expensive compared to neighboring countries (Thailand)<br>Many blamed the lack of robust public transport system which meant relying on other means of transport to reach the health care facility |
|  | Attitudes and beliefs | Feminine appearance<br>Perceived lack of need<br>Fear and anxiety | Having a beautiful smile was recognized as a sign of feminine appearance and an important aspect for attracting clients<br>Dental treatment was perceived as only required when there is pain. More emphasis is given to appearance rather than functionality<br>Dental treatment associated with fear and anxiety<br>Perceived as pain inducing |
|  |  | Fake dentists | • Ashamed of opening the mouth to show what is inside<br>• A fake dentist can provide quick fixes to their dental problems without lengthy treatment plans<br>• Online veneers and braces are easy to procure |
| Safe sexual practices relating to oral transmission of STIs | Place in the community | Stigma and discrimination<br>Socio-economic impact | • They felt neglected by their own people<br>• Lack of equal job opportunities<br>• Religious and political influences contribute to marginalization<br>• They felt 'targeted' as they stand out in the community<br>• Limited employment opportunities |
|  | Dealing with HIV/STIs | Public health care facilities<br>Awareness | • Stigma and discrimination by healthcare staff was reported to be a reason for avoiding public health care facilities even though they were freely accessible<br>• Lack of trust in the abilities of the health care provider to manage transgender health related concerns<br>• Inadequate knowledge of transgender specific aspects of health care by doctors and lack of gender affirming care facilities<br>• The participants believed that they were overloaded with information regarding HIV<br>• Fear of diagnosis prevented routine testing and regular health visits |
|  | Perception of low risk | Oral sex<br>Partner or client factors | • Lack of awareness of oral STIs<br>• Even though practiced routinely, less risk was associated with oral sex, with low potential to transmit infections |
|  | Role of support network | NGOs<br>Community | • Friendship and sisterhood are held high in value<br>• Integration into the community was crucial to seek partners or clients<br>• Community activists help with accessing testing and other health care services |

### Themes

The data was interpreted using a combination of inductive and deductive strategies. Results were represented as participants’ perspectives and researcher’s interpretations. While the in-depth interviews ascertained the participants’ personal beliefs and perspectives, the focus group discussion brought out a more general overview regarding the topic. The data analysis was guided by the interpretative lens of the Information, Motivation and Behavioural skills (IMB) theory.(36) The major themes were ‘Access to care’, ‘Attitudes and beliefs’, ‘Place in the community’, ‘Role of support network’, ‘Dealing with HIV/STI’, and ‘Perception of low risk’. Figure 1 is the thematic analysis map with major themes and minor themes that were derived from the analysis. The summary of findings from thematic analysis is summarised in table 2. Table 3 summarises theoretic analysis informed by existing literature review and data analysis from results of current study.

**Table 3:**
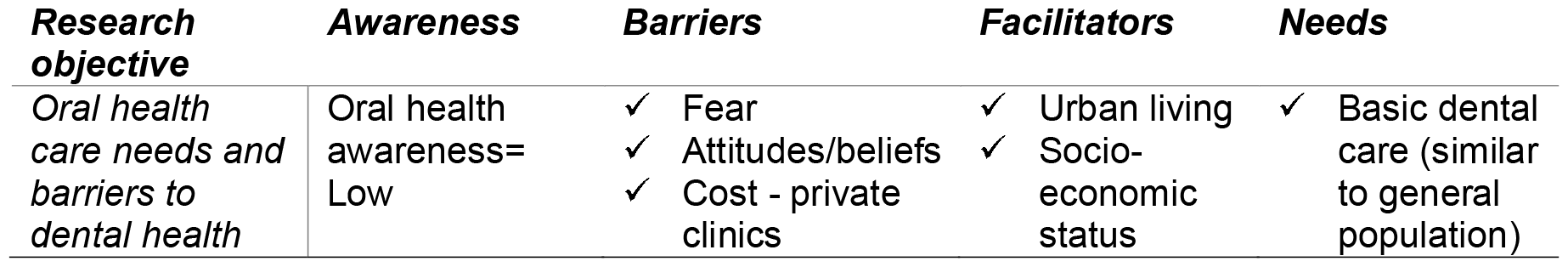
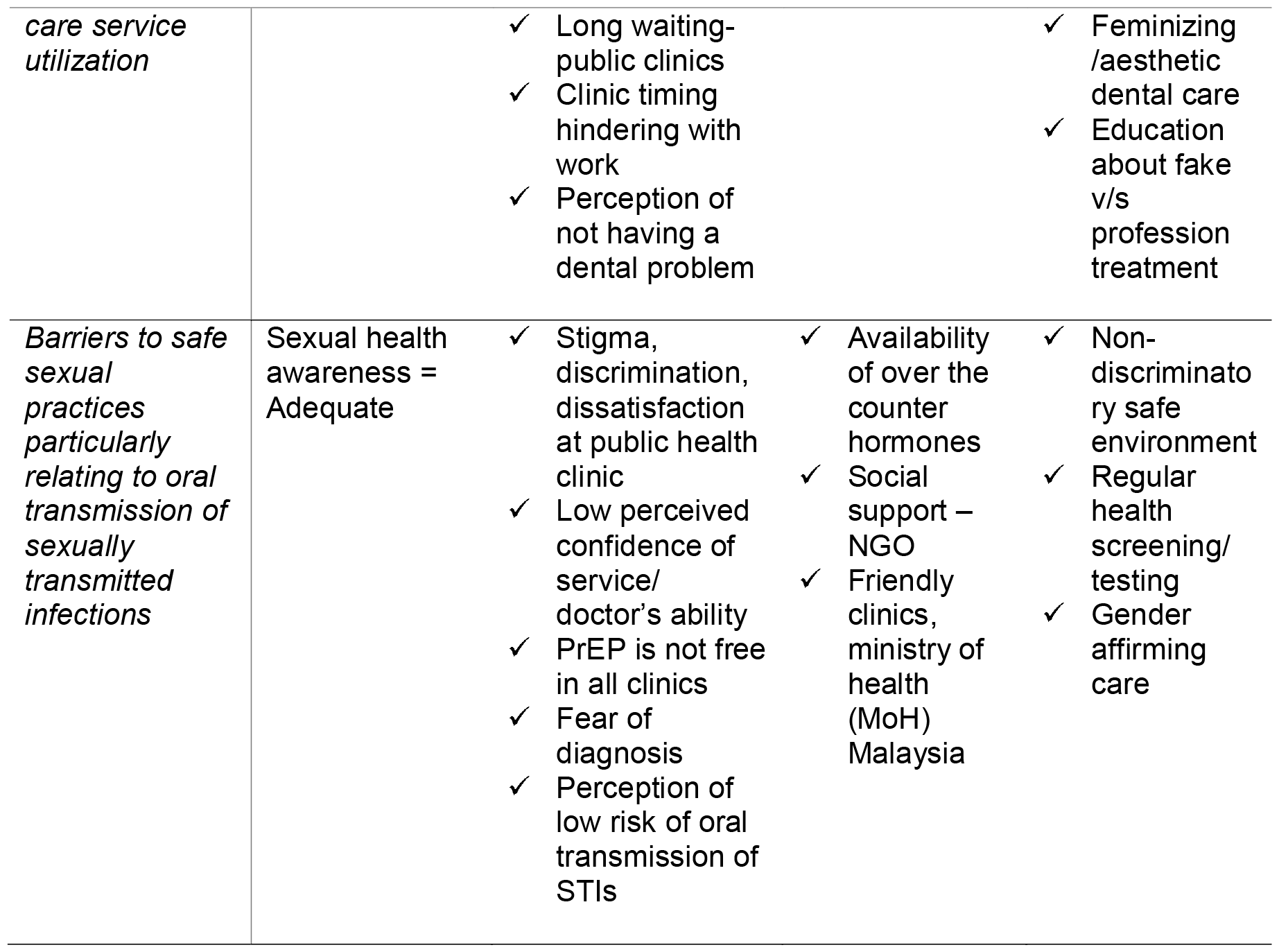
Summary of theoretic analysis informed by existing literature review and data analysis from results of current study:

**Fig 1:**
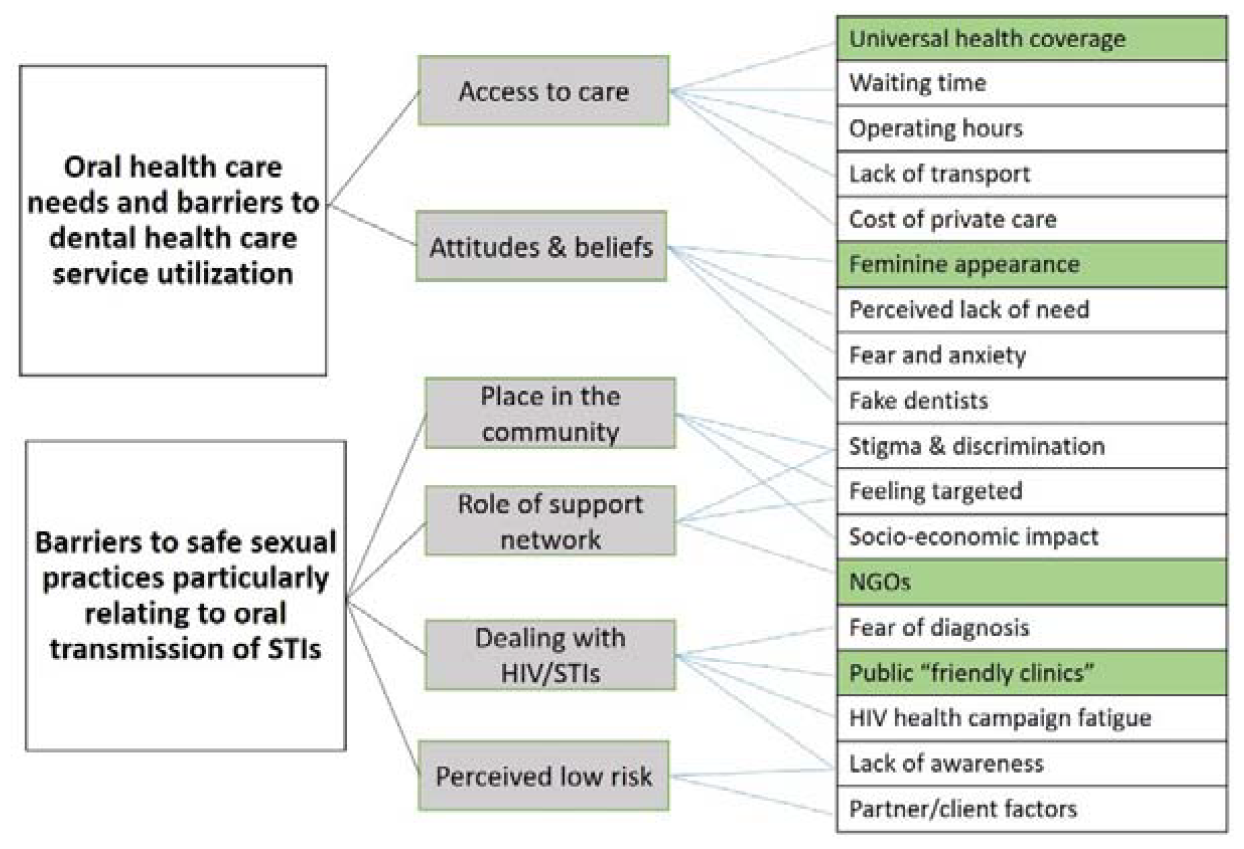
Thematic analysis map (Green boxes= enablers, White boxes= barriers)

### Access to care

The participants in general were of the opinion that health care facilities in Malaysia were available but not very accessible for transgender women. Discrimination at health care facilities was mainly from the staff and less from the doctors themselves. Another common concern was the lack gender affirming care or even the familiarity with hormone therapy by health care providers. Although the STI ‘friendly clinics” have just been set up few years ago which cater to the needs of the high risk key population, there are very few such clinics available and mostly in urban areas only.

> *“Many a times the doctors there are not familiar with specific gender related concerns of gender diverse people like me*.. *they never learnt it in Uni*..*I think*.. *(refers to University, with a ridiculed look) I went to the supposed “friendly clinic” about pain with my breast implant*.. *they were clueless*.. *or maybe did not want to treat me, because it was done in another country (referring to Thailand)*.*” Respondent 001IDI*
>
> *“I am paying taxes* .. *I can use the clinic and hospital facility for free just like anybody else*.. *I get angry just to think of that. How can the hospital staff talk to me so rudely*.. *I have been asked “how long I am planning to act like a woman” on my face*..*I was really sick and they admitted me in the male ward…. I can’t do anything*.. *no one came to see me*.. *my friends*..*how could they*.. *they are not male. I don’t want to go back to that place ever again… (Emotional and teary)-Respondent 011IDI*

In the case of dental care access, priority for visiting the dentist was found to be by far low. None of the participants interviewed has been to a dentist in the last 12 months and did not see the need to see one unless there was a specific concern. Although discrimination per se, in the dental setting was not a concern that was highlighted, discontent with clinic operating hours and a general lack of trust in the public health care facilities was opined.

> *“Oh (exclaims) the KKM clinic (Government clinic)*.. *They take so long*.. *Sometimes I wonder if they know what they are doing”. “I prefer the private dental clinic in my neighborhood*..*I can go in the evenings*.. *I cannot get up early and line up* .. *I work at night” -Respondent 019IDI*

### Attitudes and beliefs

Oral health was considered low in priority with most of the participants not having been for a dental check in the past 12 months. The need to see a dentist was only when there was a genuine problem such as pain. Home remedies and internet remedies were most often sought after. Participants were all aware that the government dental clinics were free yet preferred a private dental set up due to confidentiality concerns and overall lack of confidence in the quality of care being provided in government dental clinics.

> *“It is very easy to buy fake braces or fake veneers online*.. *very cheap also”-Respondent* 013IDI
>
> *“I think it is enough to go to dentist when you have pain only*.”-*Respondent* 006FDG
>
> *“I prefer to go to Bangkok for dental, the veneers are cheaper. Also they don’t question you about what shade you want. I like mine as white as possible*, .. *glow in the dark kind” (says it with a smile)-Respondent 0018IDI*

### Place in the community

In general there was unhappiness as they felt they were treated as second class citizens in their own country and being let down by their own people. Universally, the interview participants as well as the key informants agreed that there were lack of job opportunities for transgender women in Malaysia primarily because of their appearance. Moreover existing religious and political environment was an influencing factor that contributed to marginalization of the community.

> *“I work very hard, I have never taken medical leaves; simple*.. *in last 20 years… I want them (referring to cis-gendered persons) to see that even though I am a transwomen, I can do my job properly, so that they can know that our community is good, we can do the same jobs, don’t look at me differently!” -Respondent 013IDI*
>
> *“I make more money doing sex-work as a transwoman*.. *I was not getting a stable job, nobody wants to hire trans people*.. *But, I am happy now*.. *I make enough to get my surgery done, travel and all that!” -Respondent 018*
>
> *“I feel transwomen are targeted because we are obvious and stand out in the crowd. The others L,G, and B can hide. But the “T”, we cannot hide!” “The religious leaders spread hatred towards us*.*”-Respondent 003FDG*

### Role of support network

Support network provided by the transgender community was seen as an important aspect during the transition journey of every transgender woman. They rely on each other for emotional and social support as many of them were estranged from their families from an early age due to lack of acceptance, financial or other social concerns.

> *“It is very easy to buy hormones from the pharmacy”, but if you have to go through the proper channel, they will send you see a shrink (psychiatrist)*.. *to see if you really are a transgender in your head*.. *(rolls her eyes as she speaks)*.. *only then the doctor can prescribe you the hormones*.. *how many of us can go through that ***… (swears)… so we rely on each other and information from the internet*.*-Respondent 005IDI*
>
> *“Being a transgender woman and being Muslim is difficult*.. *The struggle is real especially in rural areas. That is why they live in big cities so that there is better community support and jobs”-Respondent 001FGD*
>
> *“It is difficult to reach the ones who default treatment (referring to HIV). We work closely with the NGOs and the community is the one that lends a helping hand for their own kind”-Respondent 002FGD*

### Dealing with HIV/STIs

Transgender women residing in urban living conditions were more aware of HIV/STI because of the awareness campaigns of the ministry of health (MoH), Malaysia. Health promotion activities are general conducted through the NGOs which employ members from the key population groups to create awareness and provide incentives. Many of the participants perceived such campaigns to be overloaded with information with little if not any new message to take home. They felt repeatedly targeted as “key population” in every health promotion activity, often being clubbed with MSMs (men who have sex with men). This was seen as a reason for “health campaign fatigue”. Also the older transwomen were of the opinion that those who newly transitioned did not pay attention to any of these campaigns and preferred to lead a risky life. For some the fear of “getting diagnosed “with a disease prevented them from seeking medical advice or even regular testing for HIV/STIs. A general lack of information regarding PrEP was also noticed.

> *“PrEP is only for HIV*.. *what about the others?(referring to other STIs)*.. *You still need protection”-Respondent 005*
>
> *“I don’t like to be mixed with the MSMs and gays. We are different, we look different*.. *that’s why the police is also after us and the KKM (government clinics) is also targeting us!”Respondent -005IDI*
>
> *“The NGOs do programs sometimes*.. *even about health, law enforcement, but the transgender women, especially the young ones* ..*don’t want to go, even they give some incentives also*.. *no they don’t care*.. *Maybe they think its not important*.*”-Respondent 016IDI*
>
> *“Only minimal training regarding transgender specific health is given to the doctors who manage the “STI-friendly clinics”, it is up to the individual doctor to build a rapport and convince the patients to come for regular follow up”-Respondent 002FGD*

### Perception of low risk

The general perception of the risk of transmission of STIs through oral sex was found to be low. Many of the transwomen having casual sex partners did not seem to think of oral sex and saliva as a potential sources of spread of infection. The use of condoms for oral sex was low due to unpleasant taste of the condom itself. Dental dams were virtually unheard of, let alone the availability or the use for them. None of the participants ever reported any oral lesions as a consequence of oral sex. The only concern was ejaculation in the oral cavity, for which the opinion was washing the mouth after the act would be sufficient to keep clean.

> *“I don’t have a partner, but I have many friends who are boys*.. *friends with benefits” “I am aware, (says reluctantly)*.. *I know condoms are needed for a blowjob( fellatio)*..*but*..*”If I know the guy is very clean*.. *I don’t ask him to wear a condom*.. *may be one out of ten…*..*hmm but*..*when you fall in love, you don’t think of condoms”-Respondent 002IDI*
>
> *“Ulcers… I don’t know, but when I sometimes involve in heavy oral sex, I have injuries in the mouth”-Respondent 008IDI*
>
> *“Saliva is antiseptic”*.. *You don’t need condoms for oral sex!*..*(looks surprised)*…*-Respondent 020IDI*

## DISCUSSION

The present study identified oral health care needs and barriers to dental service utilization in transgender women in an urban setting in Northern Malaysia. In summary, transgender women believed that it was important to have an attractive smile in order to look feminine, yet visiting a dentist was perceived as only needed when there was pain or a specific complaint. Dental treatment was associated with fear and anxiety. The perceived lack of need of dental care or the avoidance of a dental visit were findings in agreement with studies reporting the association of dental fear with experiences of discrimination and maltreatment and self-reported poor oral health.(37-39)

Barriers to utilization of public dental health services were attributed to long waiting time, unsuitable opening hours or simply the lack of transportation to reach the clinic. On the other hand private dental care was deemed expensive and hence out of reach for many. This gap in availability of suitable dental services may be a cause for turning to fake dentists or unprofessional do-it-yourself dental treatments which has become a growing concern in Malaysia. (40, 41)

As with most qualitative analyses, the interviews and focus group discussion brought out detailed understanding of the broad assumptions regarding transgender women in Malaysia. The participants who were interviewed all had a secondary school level of education or higher and belonged to urban communities. They reported experiencing marginalization, being unable to express their identities and a general discontent with their place in the society. Limited access to education, lack of job opportunities, stigma and discrimination experienced at various fronts were similar to those reported by transgender women belonging to other low-middle income countries including Malaysia (42) (4, 29) and in the global context.(43, 44) Critical factors that determined their decision to utilize public health services as well as HIV/STI clinics were influenced by socio-political environment and fear of discrimination at health care centres. Laws and policies that are discriminatory and punitive are also believed to be responsible for pushing this community away from health care services and undermining public health efforts.(4, 6) These factors along with a lack of trust in the health care system were echoed in the result of similar studies involving transgender women from within the country (2, 6, 45)and other parts of the globe.(46-49)

The barriers and facilitators to safe sexual practices particular in relation to oral transmission of STIs were also explored. The participants in the study were aware of importance of barrier protection and usage of condoms for safe sex. Although oral sex was routinely practiced, oral transmission of STIs was not particularly perceived as risky. Gaps in knowledge regarding oral STIs and risks associated with oral sex were similar to gaps in knowledge reported even in other gender diverse populations as well as heterosexual population. (23, 50-52) Even though the knowledge of spread of infection may have been heightened in general owing to the recent pandemic, spread of infection via saliva was not particularly seen as a concern. The risk of transmission of HIV through saliva and healthy oral mucosa may be none or negligible (24) but the same is not true for other viral or bacterial STIs. With growing concerns of antibacterial resistance with STIs like N. gonorrhoea, it is recommended that awareness regarding transmission of STIs via oral sexual practice be more openly discussed and more widely incorporated into public health interventions.(53) At the same time Human Papilloma Virus (HPV) transmission via oral sex and development of oral and oro-pharyngeal cancers also needs emphasis in both key populations and lay public.(52)

## CONCLUSION

Our finding from this qualitative analysis provide a perspective of a sample of transgender women living in an urban society in Northern Malaysia. They reflect the widespread sentiment within the community, feeling marginalized and discriminated against in accessing healthcare services to which they are rightfully entitled.. The finding from the interviews brought out the gaps in knowledge and provided a nuanced understanding of the needs of the community. Perceived low risk of transmission of STIs through oral sex and a general lack of awareness regarding importance of oral health was noticed along with low priority for dental care. The study also highlighted the positive role of support network in health care access and wellbeing of the population. With the awareness of existing negative oral health consequences, we would like to conclude that there is a need for a peer-reviewed, customized and culturally sensitive educational intervention along with a policy on care for this vulnerable and marginalized community.

## Limitations

The perception and views of the participants in this study were limited to that of urban living transgender women. A larger sample involving transgender women from rural areas may have varied inferences.

## Data Availability

All data produced in the present work are contained in the manuscript

## Acknowledgements

The authors would like to acknowledge the contribution of the research assistant and all those who participated in the study.

Conference presentation: Ajay Telang L, Shaik Daud H, Cotter A, Rashid A. 2069. Exploring the Barriers and Enablers of Safe Oral Sexual Practices among Transgender Women of Malaysia: A Qualitative Study on STI Transmission. InOpen Forum Infectious Diseases 2023 Dec (Vol. 10, No. Supplement_2, pp. ofad500-139). US: Oxford University Press.

## Contributors

LT and AR conceived the study. LT conducted the data collection. LT, AR and AC contributed to data analysis and interpretation. LT wrote the manuscript with supervision from AR and AC who also edited the manuscript.

## Funding

None

## Competing interests

None declared.

## Patient consent for publication

Obtained.

## Notes

### Competing Interest Statement

The authors have declared no competing interest.

### Funding Statement

This study did not receive any funding

### Author Declarations

The study received ethics approval by the University College Dublin, UCD Human Research Ethics Committee Sciences (HREC-LS) (LS-C-22-158-Telang-Cotter) and the Joint Penang Independent Ethics Committee (JPEC 22-0002)

## References

1. Poteat T, Scheim A, Xavier J, Reisner S, Baral S. Global epidemiology of HIV infection and related syndemics affecting transgender people. Journal of acquired immune deficiency syndromes (1999). 2016;72(Suppl 3):S210.

2. Gibson BA, Brown SE, Rutledge R, Wickersham JA, Kamarulzaman A, Altice FL. Gender identity, healthcare access, and risk reduction among Malaysia’s mak nyah community. Glob Public Health. 2016;11(7-8):1010–25.

3. Teh YK. The Mak Nyahs: Malaysian male to female transsexuals: Marshall Cavendish Academic; 2002.

4. Draman S, Hashi AA. Health and social challenges of LGBT: Islamic perspective. IIUM Medical Journal Malaysia. 2019;18(1).

5. Afiqah SN, Rashid A, Iguchi Y. Transition experiences of the Malay Muslim Trans women in Northern Region of Malaysia: A qualitative study. Dialogues in Health. 2022;1:100033.

6. Tan KKH, Lee KW, Cheong ZW. Current research involving LGBTQ people in Malaysia: A scoping review informed by a health equity lens. Journal of Population and Social Studies [JPSS]. 2021;29:622–43.

7. Zay Hta MK, Tam CL, Au SY, Yeoh G, Tan MM, Lee ZY, et al. Barriers and Facilitators to Professional Mental Health Help-Seeking Behavior: Perspective of Malaysian LGBT Individuals. Journal of LGBTQ Issues in Counseling. 2021;15(1):38–58.

8. Rannan-Eliya RP, Anuranga C, Manual A, Sararaks S, Jailani AS, Hamid AJ, et al. Improving Health Care Coverage, Equity, And Financial Protection Through A Hybrid System: Malaysia’s Experience. Health Aff (Millwood). 2016;35(5):838–46.

9. Lo Ying-Ru PAA. World Health Day 2018 – Lessons from Malaysia on Universal Health Coverage 2018 [Available from: https://www.who.int/malaysia/news/detail/18-04-2018-world-health-day-2018-%E2%80%93-lessons-from-malaysia-on-universal-health-coverage.

10. Ravindran TKS, Govender V. Sexual and reproductive health services in universal health coverage: a review of recent evidence from low- and middle-income countries. Sexual and Reproductive Health Matters. 2020;28(2):1779632.

11. Vijay A, Earnshaw VA, Tee YC, Pillai V, White Hughto JM, Clark K, et al. Factors associated with medical doctors’ intentions to discriminate against transgender patients in Kuala Lumpur, Malaysia. LGBT health. 2018;5(1):61–8.

12. Ngadiman S SA, Aziz MN, Yuswan F, Taib SM, Masnin H, et al. . Malaysia National Strategic Plan for Ending AIDS 2016-2030. In: HIV/STI Section DCD, Ministry of Health Malaysia, editor. 2015.

13. Suleiman A RM, Hafad FSA, Chandrasekaran S. Integrated Biological and Behavioral Surveillance Survey. In: Ministry of Health M, editor. 2017.

14. Wickersham JA, Gibson BA, Bazazi AR, Pillai V, Pedersen CJ, Meyer JP, et al. Prevalence of Human Immunodeficiency Virus and Sexually Transmitted Infections Among Cisgender and Transgender Women Sex Workers in Greater Kuala Lumpur, Malaysia: Results From a Respondent-Driven Sampling Study. Sex Transm Dis. 2017;44(11):663–70.

15. Maliya S, Zul A, Irwan M, Samsul D, Zakiah M, Rafidah H. Mak Nyahs in Malaysia: Does HIV/AIDS Knowledge Really Reduce HIV-Related Risk Behaviours? International Medical Journal Malaysia. 2018.

16. Ranjit YS, Gibson BA, Altice FL, Kamarulzaman A, Azwa I, Wickersham JA. HIV care continuum among cisgender and transgender women sex workers in Greater Kuala Lumpur, Malaysia. AIDS Care. 2021:1–7.

17. Malaysia MoH. National Strategic Plan: Ending AIDS 2016–2030. MOH Putrajaya; 2015.

18. Tharmalingam D. PrEP Initiatives in Malaysia 2023 [Available from: https://www.apcom.org/prep-initiatives-in-malaysia/.

19. Chong S, Hollingshead B, Lim S, Bourne A. A scoping review of sexual transmission related HIV research among key populations in Malaysia: implications for interventions across the HIV care cascade. Global public health. 2021;16(7):1014–27.

20. Beyrer C, Sullivan PS, Sanchez J, Dowdy D, Altman D, Trapence G, et al. A call to action for comprehensive HIV services for men who have sex with men. The Lancet. 2012;380(9839):424–38.

21. National Institute for Health and Care Excellence guideline N. Reducing sexually transmitted infections Published: 15 June 2022. 2022.

22. Garofalo R, Kuhns LM, Reisner SL, Mimiaga MJ. Behavioral Interventions to Prevent HIV Transmission and Acquisition for Transgender Women: A Critical Review. J Acquir Immune Defic Syndr. 2016;72 Suppl 3(Suppl 3):S220–5.

23. King AJ, Bilardi JE, Fairley CK, Maddaford K, Chow EP, Phillips TR. Australian sexual health service users’ perspectives on reducing the oral transmission of bacterial STIs: A qualitative study. The Journal of Sex Research. 2023:1–12.

24. Xu X, Chow EP, Ong JJ, Hoebe CJ, Williamson D, Shen M, et al. Modelling the contribution that different sexual practices involving the oropharynx and saliva have on Neisseria gonorrhoeae infections at multiple anatomical sites in men who have sex with men. Sexually Transmitted Infections. 2021;97(3):183–9.

25. Tuddenham S, Hamill MM, Ghanem KG. Diagnosis and Treatment of Sexually Transmitted Infections: A Review. Jama. 2022;327(2):161–72.

26. Saini R, Saini S, Sharma S. Oral sex, oral health and orogenital infections. J Glob Infect Dis. 2010;2(1):57–62.

27. World Health Organization. Oral Health 2024 [Available from: https://www.who.int/health-topics/oral-health#tab=tab_1

28. Kuhns LM, Hereth J, Garofalo R, Hidalgo M, Johnson AK, Schnall R, et al. A Uniquely Targeted, Mobile App-Based HIV Prevention Intervention for Young Transgender Women: Adaptation and Usability Study. J Med Internet Res. 2021;23(3):e21839.

29. Del Río-González AM, Lameiras-Fernández M, Modrakovic D, Aguayo-Romero R, Glickman C, Bowleg L, et al. Global scoping review of HIV prevention research with transgender people: Transcending from trans-subsumed to trans-centred research. J Int AIDS Soc. 2021;24(9):e25786.

30. Poteat T, Malik M, Scheim A, Elliott A. HIV Prevention Among Transgender Populations: Knowledge Gaps and Evidence for Action. Current HIV/AIDS Reports. 2017;14(4):141–52.

31. Champion VL, Skinner CS. The health belief model. Health behavior and health education: Theory, research, and practice. 2008;4:45–65.

32. Andersen RM. Revisiting the behavioral model and access to medical care: does it matter? Journal of health and social behavior. 1995:1–10.

33. Hennink M, Kaiser BN. Sample sizes for saturation in qualitative research: A systematic review of empirical tests. Soc Sci Med. 2022;292:114523.

34. Braun V, Clarke V. Using thematic analysis in psychology. Qualitative research in psychology. 2006;3(2):77–101.

35. Creswell JW, Poth CN. Qualitative inquiry and research design: Choosing among five approaches: Sage publications; 2016.

36. Fisher JD, Fisher WA. Changing AIDS-risk behavior. Psychological bulletin. 1992;111(3):455.

37. Heima M, Heaton LJ, Ng HH, Roccoforte EC. Dental fear among transgender individuals - a cross-sectional survey. Spec Care Dentist. 2017;37(5):212–22.

38. Raisin JA, Keels MA, Roberts MW, Divaris K, Jain N, Adkins DW. Barriers to oral health care for transgender and gender nonbinary populations. The Journal of the American Dental Association. 2023;154(5):384-92.e4.

39. Tharp G, Wohlford M, Shukla A. Reviewing challenges in access to oral health services among the LGBTQ+ community in Indiana and Michigan: A cross-sectional, exploratory study. PloS one. 2022;17(2):e0264271.

40. Siek JW, Salleh SM, Zaman NAK, Sinor MZ, Ahmad B. The reasons for seeking dental services from unqualified operators: A qualitative study. IIUM Journal of Orofacial and Health Sciences. 2024;5(1):30–40.

41. Nor NAM, Hassan WNW, Yusof ZYM, Makhbul MZM. Fake braces by quacks in Malaysia: a major concern. Annals of Dentistry University of Malaya. 2020;27:33–40.

42. Samuel SR, Muragaboopathy V, Patil S. Transgender HIV status, self-perceived dental care barriers, and residents’ stigma willingness to treat them in a community dental outreach program: Cross-sectional study. Special Care in Dentistry. 2018;38(5):307–12.

43. Van Gerwen OT, Jani A, Long DM, Austin EL, Musgrove K, Muzny CA. Prevalence of Sexually Transmitted Infections and Human Immunodeficiency Virus in Transgender Persons: A Systematic Review. Transgend Health. 2020;5(2):90–103.

44. Reisner SL, Poteat T, Keatley J, Cabral M, Mothopeng T, Dunham E, et al. Global health burden and needs of transgender populations: a review. The Lancet. 2016;388(10042):412–36.

45. Chong LSH, Kerklaan J, Clarke S, Kohn M, Baumgart A, Guha C, et al. Experiences and Perspectives of Transgender Youths in Accessing Health Care: A Systematic Review. JAMA Pediatrics. 2021;175(11):1159–73.

46. Matsuzaka S, Romanelli M, Hudson KD. “Render a service worthy of me”: A qualitative study of factors influencing access to LGBTQ-specific health services. SSM-Qualitative Research in Health. 2021;1:100019.

47. Pandya Ak, Redcay A. Access to health services: Barriers faced by the transgender population in India. Journal of Gay & Lesbian Mental Health. 2021;25(2):132–54.

48. Costa AB, Fontanari AMV, Catelan RF, Schwarz K, Stucky JL, da Rosa Filho HT, et al. HIV-Related Healthcare Needs and Access Barriers for Brazilian Transgender and Gender Diverse People. AIDS and Behavior. 2018;22(8):2534–42.

49. Krishnan A, Weikum D, Cravero C, Kamarulzaman A, Altice FL. Assessing mobile technology use and mHealth acceptance among HIV-positive men who have sex with men and transgender women in Malaysia. PLoS One. 2021;16(3):e0248705.

50. Sarwar G, Khan MNM, Gourab G, Irfan SD, Rahman M, Rana AM, et al. Can oral sex be performed safely among men who have sex with men (MSM) and transgender women in Bangladesh? Challenges, complexities and the way forward. Heliyon. 2023;9(4).

51. Strome A, Waselewski M, Chang T. Youths’ knowledge and perceptions of health risks associated with unprotected oral sex. The Annals of Family Medicine. 2022;20(1):72–6.

52. Brondani MA, Siqueira AB, Alves CMC. Exploring lay public and dental professional knowledge around HPV transmission via oral sex and oral cancer development. BMC Public Health. 2019;19(1):1529.

53. Chow EP, Howden BP, Walker S, Lee D, Bradshaw CS, Chen MY, et al. Antiseptic mouthwash against pharyngeal Neisseria gonorrhoeae: a randomised controlled trial and an in vitro study. Sexually transmitted infections. 2017;93(2):88–93.

